# Changing socio-demographic determinants of seasonal influenza acceptance in England during the pandemic and a framework for predicting future acceptance

**DOI:** 10.1101/2024.04.30.24305327

**Authors:** A. de Figueiredo, P. Paterson, L. Lin, S. Mounier-Jack

## Abstract

**Objective:** To assess the current level of seasonal influenza vaccine acceptance in England and establish the evolving socio-demographic determinants of seasonal influenza uptake and intent-to-vaccinate behaviours between 2020 and 2022. To provide a framework for predicting future rates of seasonal influenza uptake at sub-national scales in England.

**Design:** Two cross-sectional online surveys analysed using a Bayesian time-series multilevel model followed by poststratification to re-weight against English census data.

**Setting:** England, September 2020 to July 2022.

**Participants:** 28,748 English adults, 18 years of age and older.

**Main outcome measures:** Three response variables: whether an individual was offered a seasonal influenza vaccine on the NHS in the last 12 months, whether this offer was accepted, and whether they would accept a seasonal influenza vaccine in the next 12 months.

**Results:** In the 2021-22 flu season, 56.3% of adults in England were offered the seasonal influenza vaccine, marking a significant increase of 10.7 percentage points compared to the 2019-20 season, due to the expanded rollout of the programme during the pandemic. Acceptance of the seasonal influenza vaccine saw a marked rise during this period across most age groups and particularly among individuals aged 50 and over. However, intentions to accept the vaccine in the next 12 months showed a slight decline across the English adult population between 2020 and 2022 surveys. Socio-demographic traits play a significant role in shaping vaccine behaviours, with age, gender, education, ethnicity, and religion influencing the likelihood of being offered the vaccine, accepting it when offered, and intending to receive future vaccinations. Noteworthy differences were observed across demographic groups, particularly between Black/Black British and White respondents, although gaps in acceptance between socio-demographic groups in the 65-and-over cohort were narrower than for population as a whole. Of particular concern is waning intent-to-accept behaviours among Asian and Asian British groups, as well as Hindus and Punjabi speakers. Regional disparities also emerged, with estimates for flu vaccine receipt and future intent to accept a flu vaccine relatively low in London, especially inner London. Predictions of flu vaccine uptake generated from 2022 data correlated well with the observed UK Health Security Agency-reported uptake in the subsequent 2022-23 flu season, highlighting the ability of multilevel regression and poststratification to accurately capture future intentions.

**Conclusions:** The findings underscore the significant progress made in increasing seasonal influenza vaccine uptake among adults in England during the 2021-22 flu season. Despite these improvements, disparities persist across socio-demographic groups, spotlighting the need for targeted interventions to address uptake inequity. The slight decline in intention to accept the vaccine in the general adult population warrants attention. Furthermore, regional disparities emphasise the importance of tailoring vaccination strategies to address specific geographical contexts. The strong correlation between predicted and observed vaccine uptake and observed indicates the utility of predictive modelling in informing future vaccination behaviours and public health interventions. Overall, these findings provide valuable insights for policymakers and public health practitioners to enhance influenza vaccination efforts and mitigate the burden of influenza-related illness in England.

## Introduction

The arrival of Covid-19 in the UK in early 2020 posed considerable challenges to the National Health Service (NHS), which grappled with mitigating the overall impact of the pandemic. The immunisation system fought on multiple fronts: maintaining access to and delivery of routine immunisations; the rapid distribution of novel Covid-19 vaccines; associated communication and outreach drives; and the delivery of seasonal influenza vaccines to at-risk groups. These efforts were complicated by social distancing measures and shielding of at-risk groups, as well as the possible burden of a “twindemic”. The risk of co-infection with SARS-CoV-2 and influenza was a notable concern, with co-infection carrying a six-fold increased risk of death compared to infection with neither pathogen [1]. These additional risks led to the expansion of the seasonal influenza vaccination programme in the winter of 2020 to include adults aged 50 to 64 as well as household members of patients shielding from Covid-19 [2]. In the face of these challenges, seasonal influenza uptake among over 65s increased by nearly eight percentage points between the 2019-20 and 2020-21 flu seasons (from 72.4 to 80.9) and a further two points in the 2021-22 campaign (from 80.9 to 82.9), before declining slightly to 79.9% in the 2022-23 season [3]. Between the 2020-21 and 2021-22 seasons, coverage among the expanded cohort of 50-to 64-year-olds increased from 45.2 to 52.5%, before declining to 28.6% in 2022-23, with the expanded offer not made from 2023-24 onwards [2, 3].

Despite these successes, the pandemic has brought pre-existing challenges into focus and has given rise to new ones. Inequities in vaccination behaviours, notably with respect to ethnicity, remain for a range of immunisation programmes [4, 5, 6], including the seasonal influenza programme in which large disparities in uptake exist between Black or Black British and White individuals [7] as well as with the Covid-19 vaccine, in which this ethnicity gap appears to be widening [8]. The impact of multiple pandemic factors, such as misinformation and pandemic policies, on public attitudes towards vaccination remain ambiguous, though emerging evidence points to younger groups exhibiting lower levels of vaccine confidence than before the pandemic. Young adults in the EU, for example, exhibit lower levels of vaccine confidence in 2022 compared to before the pandemic [9], and were more likely to report Covid-19 vaccine refusal if proof-of-vaccination requirements were to be implemented [10, 11]. Recent evidence from the UK also suggests that younger groups seem less robust at discerning real from fake news [12]. Though, it should be cautioned that direct causal evidence between specific pandemic factors and potential spillover effects onto other vaccines has not been established and the impact of the pandemic on attitudes to immunisation remains an open question.

While identifying causal links between pandemic factors and attitudes towards the seasonal influenza vaccine is beyond the scope of this study, we answer core questions about the landscape of seasonal influenza vaccine in the UK during the pandemic. Recent studies have explored a wide arrange of putative drivers of seasonal influenza uptake in the UK, including socio-demographics, psychological factors, risk perceptions, and previous experiences [13, 14]. In this study, we focus on exploring the socio-demographic determinants of seasonal influenza vaccination behaviours in two surveys conducted in 2020 and 2022. Specifically, we investigate the extent to which different groups report being offered a seasonal influenza vaccine, their acceptance (given the vaccine was offered), and future intent to accept a flu vaccine. Further, we establish whether socio-demographic groups express lower sentiments around the seasonal influenza vaccine in 2022 compared to prior to the introduction of the Covid-19 vaccine in late 2020. We also identify whether future rates of seasonal influenza uptake can be predicted at national and sub-national via large-scale survey data collection using multilevel regression and poststratification using English census micro-data records to re-weight individual responses. We do this by asking about future vaccination intent and validate findings against observed sub-national seasonal influenza uptake in the 2022/23 winter campaign, which corresponds to intentions stated in the 2022 survey.

Our study provides new insights into the changing landscape of seasonal influenza behaviours and inequities in England and considers a framework for forecasting future rates of seasonal influenza immunisation. We conclude our study by providing an overview of possible explanations for changes in observed behaviours.

## Methods

### Data

We use data from two large nationally representative cross-sectional surveys conducted in 2020 and 2022 among adult residents of the UK. The first of these surveys was conducted in September and October 2020 and comprised 16,906 individual surveys after data quality checks were performed. The second survey was conducted in June and July 2022 with 17,199 interviews after quality control checks. In both surveys, quotas were set for sex, age, and sub-national region according to marginal frequencies in English census data [15]. We note, however, that we deploy a further poststratification re-weighting step to extend this marginal matching on three variables to a matching on joint distributions for a wider set of socio-demographic variables via poststratification – see Multilevel time-series model and poststratification below.

We limit our analysis to residents of England for two reasons. Firstly, census micro-data records in Scotland, Wales, and Northern Ireland encode some census demographic variables differently than in England. Aligning groups would necessitate removing some important demographic groups in England, limiting the ability to identify trends in some sociodemographic groups. Secondly, indices of multiple deprivation exist for England at regional levels [16]; these indices permit an exploration of relationship between deprivation and our three response variables. This exclusion criterion reduces the respective sample sizes to 14,222 (2020 survey) and 14,526 (2022 survey) after respondents who do not reside in England are removed.

We focus on three response variables: whether a respondent has been offered a seasonal influenza vaccine on the NHS in the past 12 months; whether the respondent accepted this offer; and whether a respondent intends to accept a seasonal influenza vaccine in the next 12 months. We caution that the precise question wording of this final item differs slightly between survey waves (table 1). In the 2020 survey, the preceding 12 months would capture the 2019-20 flu season, while the following 12 months would capture the 2020-21 season. In the 2022 survey, the preceding 12 months would capture the 2021-22 flu season, while the following 12 months would capture the 2022-23 season.

**Table 1:**
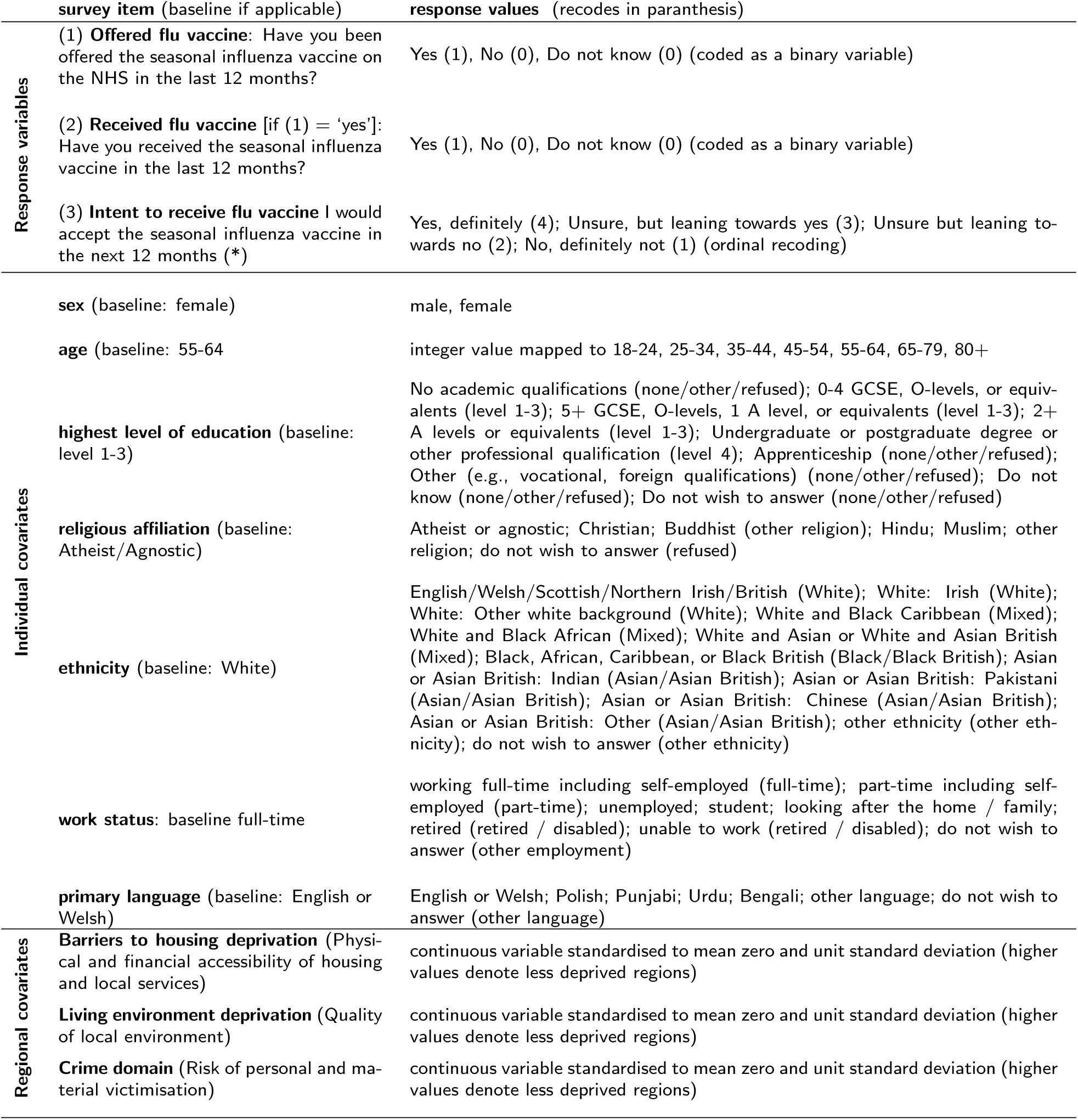
Data summary. Description of the three study response variables (being offered a flu vaccine, receiving the flu vaccine given it was offered, and intent to accept a flu vaccine in the next 12 months), as well as individual- and regional-level covariate data. (*) We note that the precise wording to response varibale (3) varies between both surveys. In 2020, respondents were asked to respond to the statement “I would accept the seasonal influenza vaccine in the next 12 months to protect myself against flu”. In 2022, respondents were asked “I would accept the seasonal influenza vaccine in the next 12 months if it was recommended to me.”

Covariate information is considered at both the individual and regional level. At the individual level, socio-demographic data are collected on respondents’ sex, age, highest level of educational achievement, religion, ethnicity, and employment status. Individuals’ outer-post code was also collected, which was used to assign each participant to one of 33 international territorial level (ITL) codes representing a sub-national classification of England [17]. These individual-level covariates were included in the survey design to both explore the major socio-demographic drivers of our response variables and to permit representative estimates of each response variable at national and sub-national levels via multilevel regression and poststratification (see Methods: Multilevel time-series model and poststratification). The 2011 UK microdata census records [15] were used for poststratification, which requires that encoding of survey covariate data matches that in the census. Although post-stratification provides an additional level robustness to marginal quotas used in data collection, we observe good agreement between survey and census demographic proportions for each demographic variable and sub-national region (figure 1).

**Figure 1:**
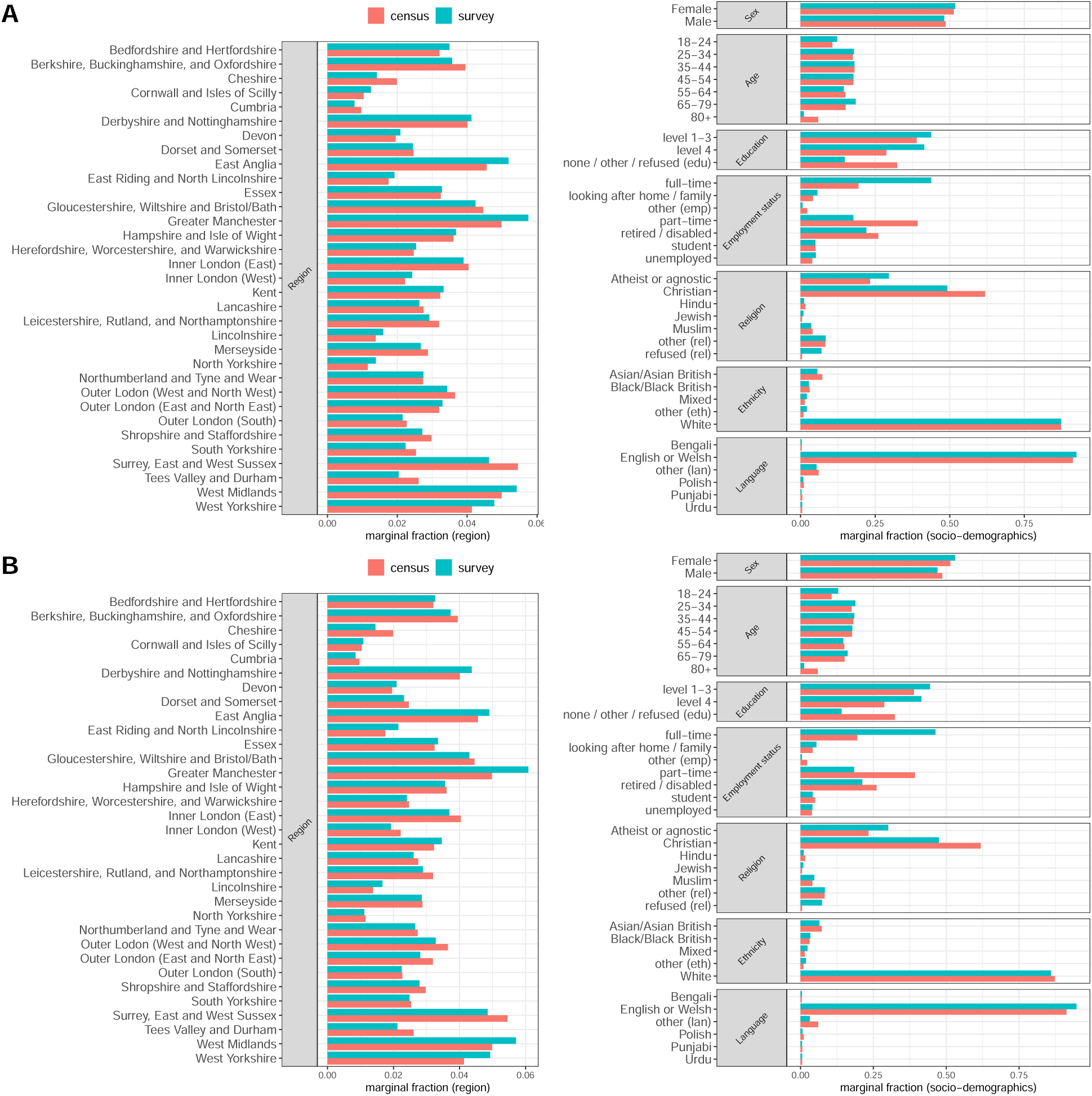
Comparison of socio-demographic and regional variable counts between survey and UK census data for the 2020 (A) and 2022 surveys (B). Within each socio-demographic category, the frequency of variable responses are shown for survey and census data.

The 2019 English indices of multiple deprivation (IMD) [16] are used as regional covariates to explain variability in regional level outcomes, with the mean index taken for all lower authority areas in each of the 33 ITL regions. IMDs measure deprivation in different areas of England and were developed by the Ministry of Housing, Communities & Local Government (formerly the Department for Communities and Local Government). The IMD is composed of several domains of deprivation, each measured using various indicators. The domains include income deprivation (the proportion of the population experiencing income deprivation and based on factors such as the proportion of the population on income-related benefits or low incomes); employment deprivation (the working-age population who are unemployed or longterm sick, and the proportion of young people not in employment, education, or training); health deprivation and disability (life expectancy, disability, and the general health of the population); education, skills, and training deprivation (indicators such as the proportion of the population with no qualifications and levels of school attainment); crime (various aspects of crime rates, including both reported and recorded crime levels); barriers to housing (indicators related to housing quality, access to services such as healthcare and education, and the quality of the local environment); and living environment deprivation (aspects such as air quality, traffic levels, and access to green spaces). While we initially considered all domains of multiple deprivation, only three domains were selected for the analysis (crime, housing, and environment) as the introduction of any additional domain induced at least one pairwise Pearson correlation greater than 0.7, which led to multi-collinearity in the Bayesian multilevel model as detected through poor convergence of posterior samples. All response and covariate data are described in table 1 along with details of variable recoding. There were no missing data on response variables and while some respondents did not provide an answer to specific covariates, we created a ‘missing’ category for these responses to avoid the loss of missing data and enable matching with census micro-data records.

### Multilevel time-series model

A multilevel model followed by poststratification (MRP) is implemented to provide national and sub-national estimates of our three response variables as well as to yield their socio-demographic drivers in 2020 and 2022. The multilevel model we deploy closely follows a recent multilevel model for cross-sectional time-series data by Kusano and Kemmelmeier [18], however we deploy a Bayesian approach. This MRP model is described below.

Let 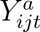 denote the response by individual *i* residing in region *j* at time *t* (where *t* = 1 is the 2020 survey and *t* = 2 is the 2022 survey) for response variable *a*. Here, *a* = 1, 2, or 3 indexes each of our response variables: *a* = 1 corresponds to whether an individual has been offered a flu vaccine in the last 12 months; *a* = 2 corresponds to whether this offer was accepted, and the flu vaccine was received; and *a* = 3 relates to intent to accept a seasonal influenza vaccine in the next 12 months. The first two variables contain two possible responses (table 1) and therefore 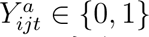 for *a* = 1, 2. The third question is answered on an ordinal scale and so 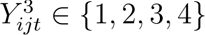 (table 1).

For *a* = 1, 2 we model the likelihood and link function as,

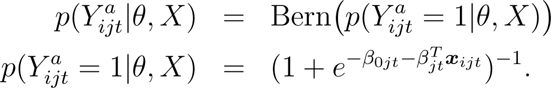

For *a* = 3, we use ordinal logistic regression with the proportional odds assumption with a threshold parameter *τ_k_*(*k ∈ {*1, 2, 3*}*),

For *a* = 1, 2 we model the likelihood and link function as,

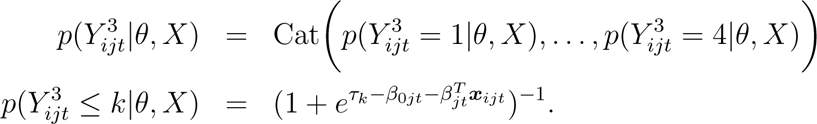

In both models, *X ∈* ℝ*^N×P^* and ***x****_ijt_* is a row of *X* containing covariate data for individual *i* at time *t*. Hierarchical priors are placed over the regression parameters to account for temporal and spatial variability, 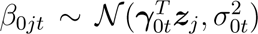 and 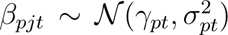 with *γ*_0_*_qt_ ∼ N* (*δ*_0_*_q_, σ̃*_0_) (for *q* = 1, 2, 3) and *γ_pt_ ∼ N* (*δ_p_, σ̃_p_*) for *p* = 1*, . . . , P* and where ***z****_j_ ∈ ℝ*^3^ are the regional covariates for the three deprivation scores. Semi-informative priors are placed on *δ*_0_*_q_* and *δ_p_*: *δ*_0_*_q_, δ_p_ ∼ N* (0, 2.5^2^). Half-normal priors are placed on the hierarchical variance components, 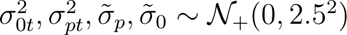 in line with recommendations [19, 20].

### Post-stratification

We take the following post-stratification re-weighting approach. Posterior predictive distributions are generated for each unique individual in the English census micro-data records. An MRP estimate can then be obtained via

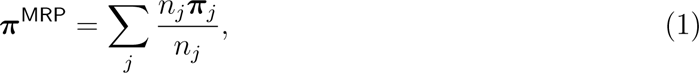

where ***π****_j_ ∈ ℝ^S^* are posterior samples for a specific unique cell *j*, and *n_j_* is the number of people in that cell in the census micro-data. If we wish to generate a national estimate, then *j* will index through each unique combination of all socio-demographic variables in the census micro-data. However, if we want to generate an estimate within a sub-group (such as a region or specific socio-demographic group), then *j* indexes over all unique cells relevant to that sub-group. For instance, if we wanted to compute an estimate for a specific region, we would find all unique socio-demographic cells in the census micro-data for that region and weight their posterior probability by the frequency of occurrence.

### Estimation

Posterior distributions are obtained via Gibbs sampling using the package rjags in R [21] using 5,000 simulations after successful model burn-in. Convergence of posterior samples are assessed using the Gelman-Rubin diagnostic [22]. The scale reduction factor satisfies *R̂ ≤* 1.05 for all parameters across all models. Posterior predictive samples are obtained via Monte Carlo integration using 100 posterior draws.

### Ethical approval

Survey data collection from 2020 was approved by the Imperial College Research Ethics Committee on 24 July 2020 with reference 20IC6133. Survey data collection from 2022 was approved on 31 March 2022 by the London School of Hygiene & Tropical Medicine’s Ethics Committee with reference 26854.

## Results

### National estimates

Across England, we estimate that 56.3% of adults (18+) (95% credible interval [CrI], 55.3 to 57.3) were offered the seasonal influenza vaccine on the NHS in the last 12 months in the 2022 survey (corresponding to the 2021-22 flu season) compared to 45.6% (CrI 44.7 to 46.5) in the 2020 survey (2019-20 season). These values represent an increase of 10.7 (9.4 to 11.8) percentage points [pp] for the adult population (table 2). There was a slight decrease in the percentage of those aged 65-and-over (65+) who report being offered a seasonal influenza vaccine in the last 12 months, however, decreasing from 89.5% (CrI 88.2 to 91.0) in the 2019-20 season to 87.2% (CrI 85.8 to 88.8) in 2021-22, a fall of 2.3pp (CrI 0.3 to 4.2) (table 2).

**Table 2:** National-level multilevel regression and poststratification estimates for the three response variables for the English adult population (18+) and the population aged 65 and over (65+) Estimates are provided for the 2020 and 2022 surveys which correspond to the 2019-20 and 2021-22 flu seasons for the first two response variables (offered a flu vaccine in the last 12 months and received a flu vaccine in the last 12 months), respectively. Estimates are provided with associated 95% credible interval.

|  | cohort | response | 2020 (%) | 2022 (%) | change (pp) |
| --- | --- | --- | --- | --- | --- |
| **Offered a flu vaccine last 12 months** | 18+ | Yes | 45.6 (44.7, 46.5) | 56.3 (55.3, 57.3) | +10.7 (9.4, 11.8) |
|  |  | No | 54.4 (53.5, 55.3) | 43.7 (42.7, 44.7) | -10.7 (-11.8, -9.4) |
|  | 65+ | Yes | 89.5 (88.2, 91.0) | 87.2 (85.8, 88.8) | -2.3 (-4.2, -0.3) |
|  |  | No | 10.5 (9.0, 11.8) | 12.8 (11.1, 14.2) | +2.3 (0.3, 4.2) |
| **Received flu vaccine last 12 months** | 18+ | Yes | 62.9 (61.4, 64.5) | 71.7 (70.4, 73.2) | +8.8 (6.7, 10.5) |
|  |  | No | 37.1 (35.5, 38.6) | 28.3 (26.8, 29.6) | -8.8 (-10.5, -6.7) |
|  | 65+ | Yes | 72.6 (69.5, 75.9) | 86.6 (84.7, 88.2) | +14.0 (10.0, 17.2) |
|  |  | No | 27.4 (24.1, 30.4) | 13.4 (11.7, 15.3) | -14.0 (-17.2, -10.0) |
| **Would accept flu vaccine next 12 months** | 18+ | Yes, definitely | 58.5 (57.6, 59.4) | 57.5 (56.4, 58.6) | -0.9 (-2.0, 0.5) |
|  |  | Unsure, lean yes | 17.9 (17.2, 18.4) | 17.3 (16.9, 17.9) | -0.5 (-0.9, -0.2) |
|  |  | Unsure, lean no | 11.6 (11.3, 12.0) | 11.9 (11.5, 12.3) | +0.3 (-0.1, 0.6) |
|  |  | No, definitely not | 12.0 (11.5, 12.5) | 13.2 (12.5, 13.8) | +1.2 (0.6, 1.8) |
|  | 65+ | Yes, definitely | 80.2 (78.4, 81.9) | 84.4 (82.9, 85.9) | +4.2 (1.8, 6.6) |
|  |  | Unsure, lean yes | 10.4 (9.5, 11.3) | 8.3 (7.6, 9.0) | -2.1 (-3.2, -1.0) |
|  |  | Unsure, lean no | 5.1 (4.6, 5.6) | 4.0 (3.6, 4.4) | -1.1 (-1.8, -0.4) |
|  |  | No, definitely not | 4.2 (3.8, 4.6) | 3.3 (2.9, 3.7) | -0.1 (-0.2, -0.0) |

We find an 8.8pp (CrI 6.7 to 10.5) increase in the proportion of adults (18+) who report receiving a flu vaccine in the 2021-22 flu season compared to 2019-20 (table 2). Despite the slight decrease in 65-and-overs reporting that they had been offered a flu vaccine, we find a large increase in the percentage of the 65+ cohort receiving the flu vaccine, given that it was offered, from 72.6% (CrI 69.5 to 75.9) to 86.6% (CrI 84.7 to 88.2), an increase of 14.0pp (CrI 10.0 to 17.2).

In the 18+ cohort, we find a 0.9pp (CrI 0.5 to 2.0) decrease in respondents who state that they would ‘yes, definitely’ accept a flu vaccine in the next 12 months in our 2022 survey compared to 2020 (2). In the 65+ cohort, however, we find a 4.2pp (CrI 1.8 to 6.6) increase in this value (table 2).

### Sub-national estimates

For the 18+ cohort, there are broad increases across England in the percentage of respondents being offered a flu vaccine (figure 2A-C) and accepting the flu vaccine, given it was offered to them (figure 2D-F) in 2022 compared to 2020 (corresponding to the 2021-22 and 2019-20 flu seasons respectively). However, we detect slight decreases across most of England in those intending to accept a flu vaccine in the next 12 months (figure 2 G-I).

**Figure 2:**
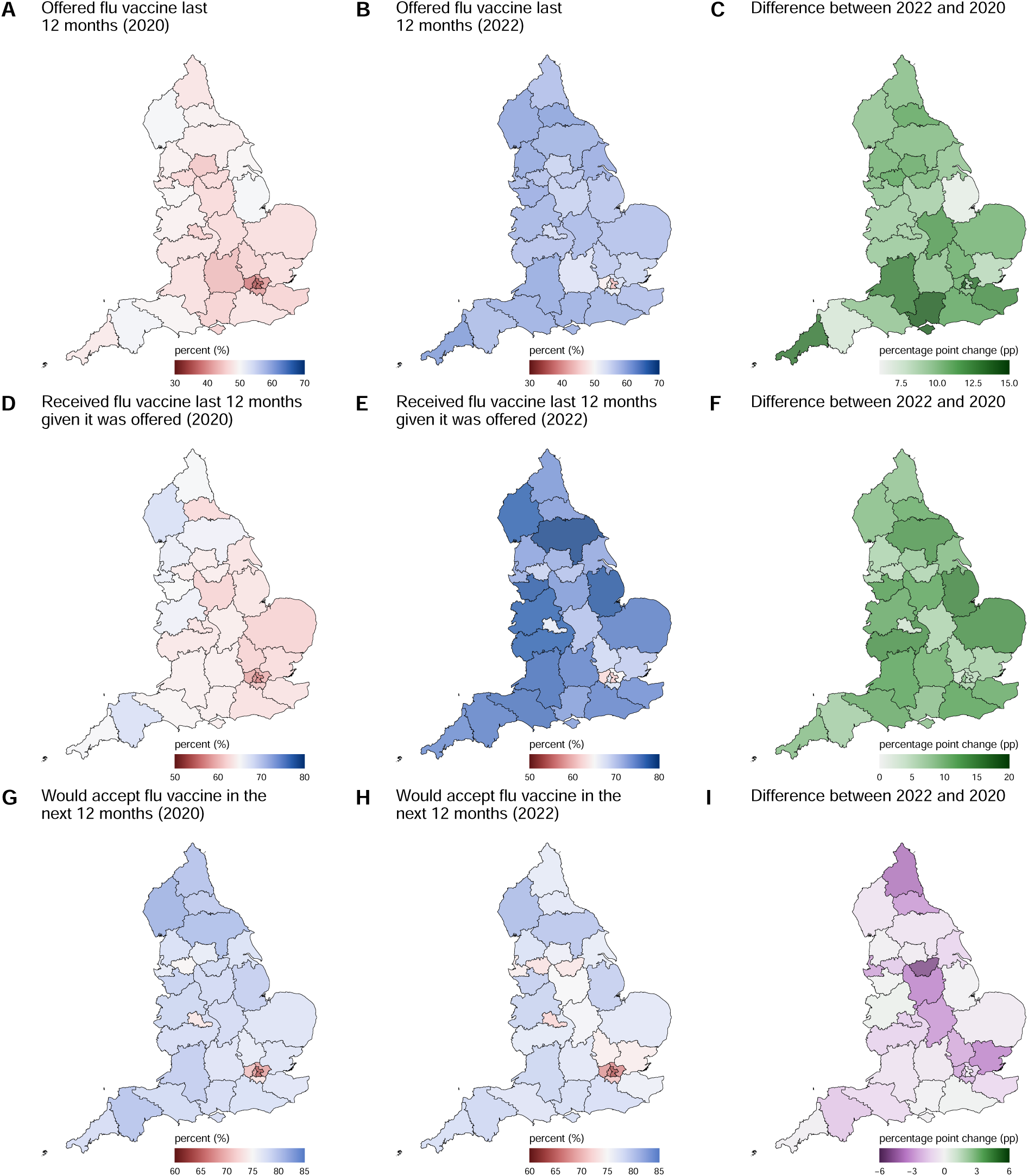
Sub-national mapping of seasonal influenza vaccination behaviours for the adult population (18+)

In the 2020 and 2022 surveys, we find that Inner London (East) had the lowest proportion of adults 18-and-over reporting that they had been offered the flu vaccine (34.8% and 43.8%, respectively). Devon had the highest in 2020, while Cornwall and Isles of Scilly had the highest in 2022 (50.6% and 61.1%, respectively, see figure 2A and B). Inner London (West) reports the largest increase in the percentage of adults being offered a flu vaccine between 2020 and 2022 (14.5), while Lincolnshire (6.6) reports the lowest increase (figure 2C). We find that Inner London (East) had the lowest proportion of adults reporting that they had received the seasonal influenza vaccine (given it was offered) in both survey years – 56.3% in 2020 (2019-20 flu season) and 61.6% in 2022 (2021-22 season). Cumbria had the highest proportion of adults reporting flu vaccine acceptance in 2020 (2019-20 season), while North Yorkshire had the highest in 2022 (2021-22 season) with values of 67.9% and 78.8%, respectively (figure 2D and E). West Midlands reported the smallest increase in the percentage of adults receiving a flu vaccine (3.6pp), while Lincolnshire (14.5pp) the largest (figure 2F). London also contained the lowest rates of the adult population (18+) reporting that they would accept a seasonal influenza vaccine in the next 12 months in both 2020 and 2022 surveys (figure 2G and H).

Among the 65-and-over cohort, London reported lower flu uptake levels (given the vaccine was offered) than the rest of the country in both 2020 and 2022 surveys and we find a striking difference in future intent to accept a seasonal influenza vaccine in both 2020 and 2022 surveys among 65-and-overs who reside in London, particularly inner London (figure 3G-H). However, across the whole of England – including all regions of London – intent to accept a flu vaccine was reported to be higher in 2022 than in 2020 (figure 2I).

**Figure 3:**
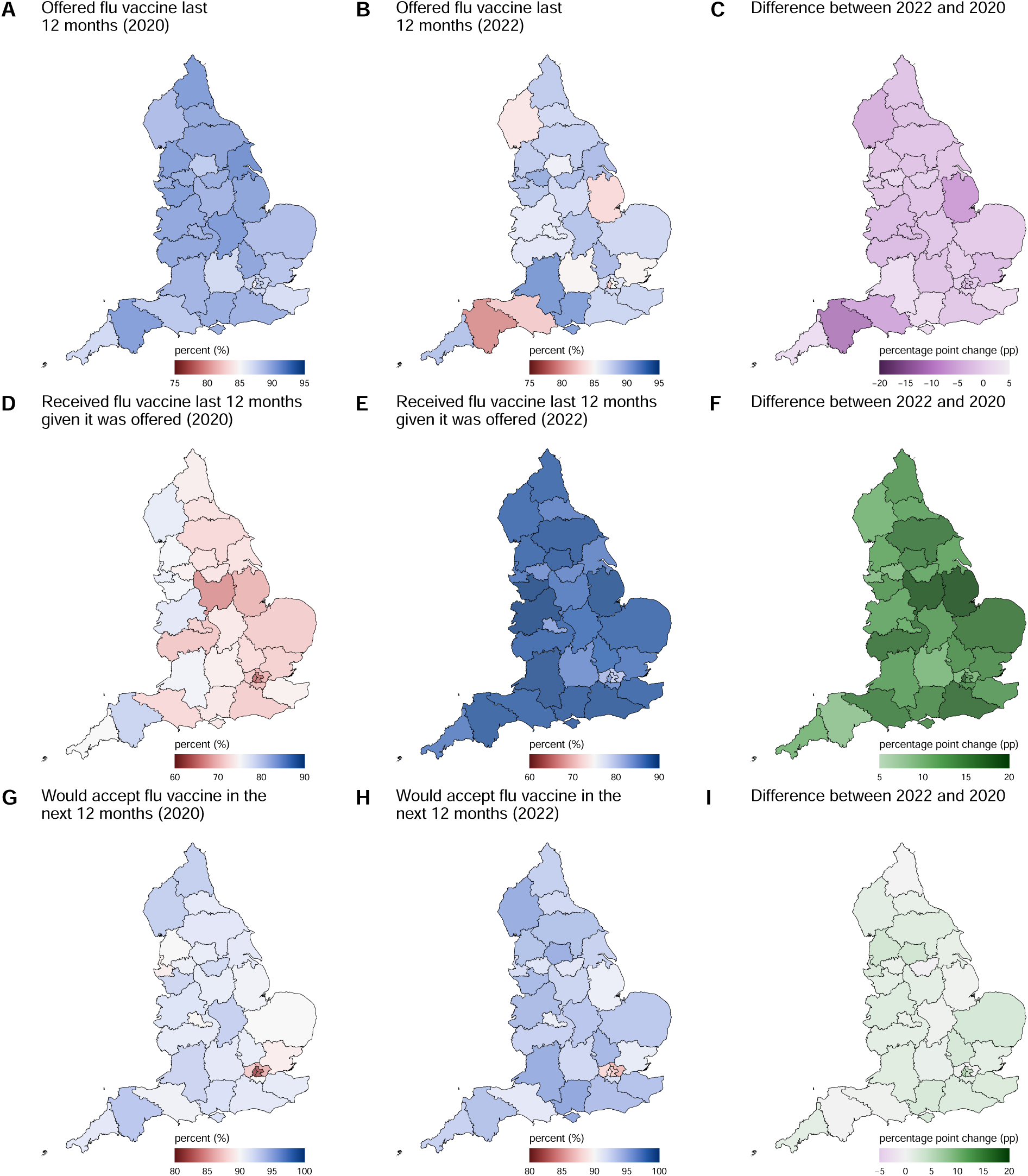
Sub-national mapping of seasonal influenza vaccination behaviours for 65-and-overs (65+)

### Individual socio-demographic determinants

Associations between individual and regional covariates and each of our three response variables obtained through our multilevel time-series model are shown in figure 4. Individual associations are presented as *adjusted* odds ratios 4A. MRP estimates for each socio-demographic group within each question and for both time points are provided in figures A.1 and A.2. These estimates are obtained using equation 1 and do not, therefore, depend on a baseline group.

**Figure 4:**
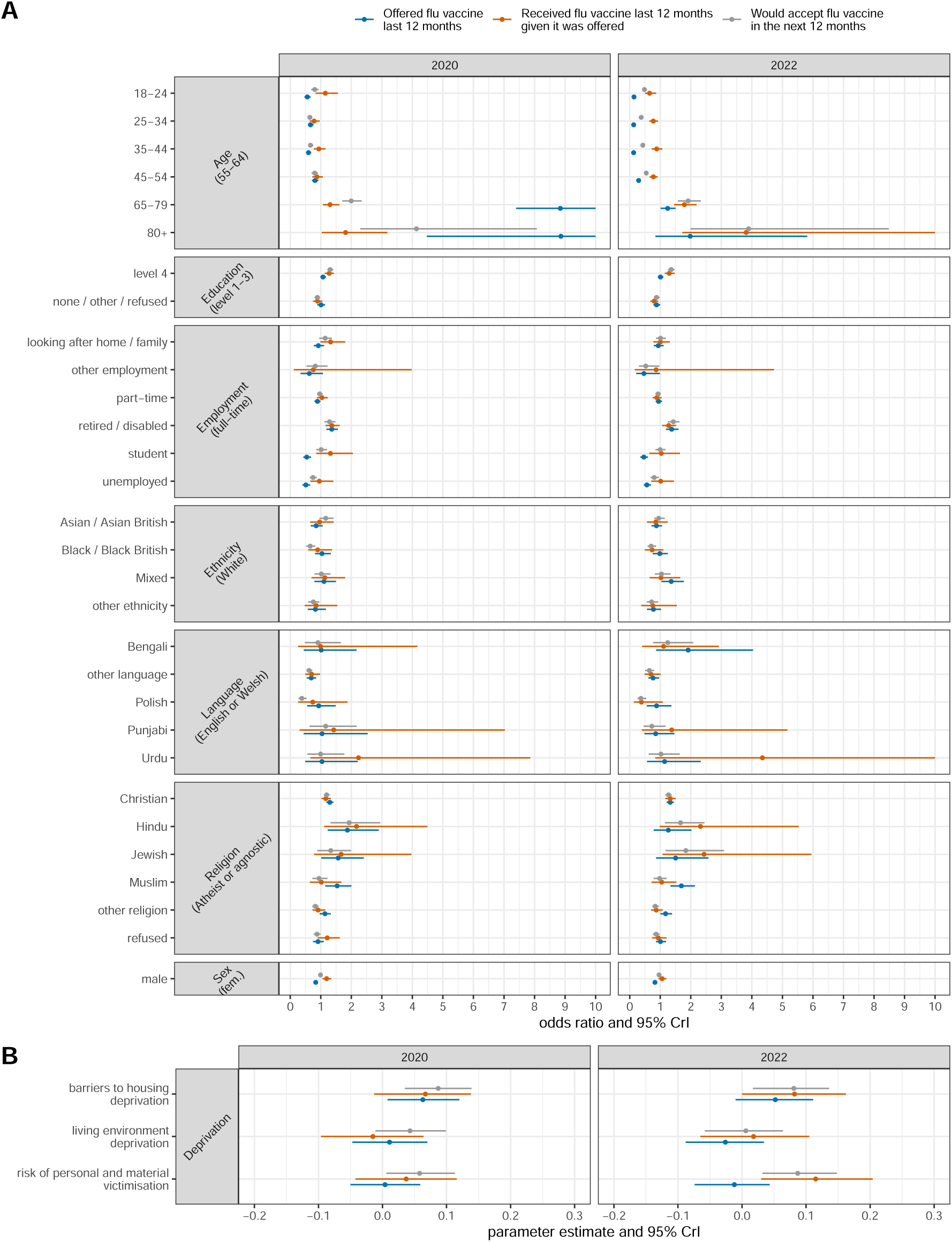
Associations between our three response variables and individual- and regional-level covariates in the 2020 (left panels) and 2022 surveys (right panels) Individual-level associations are provided as odds ratios with associated 95% credible intervals. Regional-level associations are reported as the coefficient *γ*_0_*_t_*. Odds ratios are capped at 10 to assist legibility.

The results of the multilevel logistic regression revealed that males had lower odds of reporting being offered a flu vaccine compared to females in both 2020 (odds ratio [OR] 0.83, 95% credible interval [CrI] 0.77 to 0.90) and 2022 (OR 0.83, CrI 0.76 to 0.90) (figure 4A). Individuals with lower levels of education also had lower odds of being offered the vaccine in 2022 (OR 0.88, CrI 0.70 to 1.00), while those of mixed ethnicity had higher odds in 2022 than Whites (OR 1.36, CrI 1.04 to 1.77) (figure 4A). All employment types except for retirees or disabled individuals were more likely to report being offered the flu vaccine in the last 12 months in both 2020 and 2022 surveys compared to full-time workers (figure 4A). Hindus had higher odds of being offered the vaccine in 2020 (OR 1.87, CrI 1.23 to 2.90), and Christians and Muslims had higher odds in both years compared to atheists or agnostics (Christian, 2020: OR 1.29, CrI 1.18 to 1.41; 2022: OR 1.32, CrI 1.21 to 1.44; Muslim, 2022: OR 1.53, CrI 1.15 to 2.00, 2022 OR 1.69, CrI 1.33 to 2.13) (figure 4A).

Younger groups, lower education levels, and speakers of ‘other’ languages typically had lower odds of receiving the flu vaccine in both 2020 and 2022 surveys compared to their respective baseline groups (figure 4A). We also find that Hindu and Jewish respondents had higher rates of acceptance than atheists/agnostics (figure 4A).

Regarding intent to receive a seasonal influenza vaccine in the next 12 months, we again found lower intent among younger age groups and lower education levels in both years. Black/Black British respondents were found to be much less likely than Whites to report an intent to accept a seasonal influenza vaccine in both 2020 and 2022 surveys, which correspond to the 2020-21 and 2022-23 flu seasons (2020: OR 0.65, CrI 0.52 to 0.82; 2022: OR 0.70, CrI 0.58 to 0.86). ‘Other’ ethnicity, Polish speakers, respondents who speak an ‘other’ language, and respondents who practice an ‘other’ religion were also found to have much less intent to vaccinate in the next 12 months compared to their respective baseline groups (‘other’ ethnicity, 2020: OR 0.75, CrI 0.59 to 0.95, 2022: OR 0.72, CrI 0.56 to 0.93; Polish speakers, 2020: OR 0.38, CrI 0.27 to 0.53, 2022: OR 0.37, CrI 0.25 to 0.54; ‘other language’, 2020: OR 0.61, CrI 0.53 to 0.72, 2020: OR 0.64, CrI 0.51 to 0.80; ‘other’ religion, 2020: OR 0.82, CrI 0.72 to 0.92 and 0.83, 2022: OR 0.74 to 0.95). Individuals who did not provide religious information were also less likely than atheists/agnostics to state an intent to receive the flu vaccine in the next 12 months (2020: OR 0.87, CrI 0.77 to 0.99; 2022: OR 0.86, CrI 0.76 to 0.98).

### Regional socio-demographic determinants

The deprivation scores most consistently associated with regional-level variation in the three response variables are barriers to housing and risk of personal and material deprivation. Higher levels of housing deprivation are associated with a lower chance of both being offered a flu vaccine in both surveys and receiving the vaccine given it was offered (2022 survey only) (figure 4B). Regions with higher levels of personal and material risk deprivation are associated with lower rates of being offered the flu vaccine (both surveys), while higher levels of personal and material risk deprivation is associated with lower flu vaccine receipt in the 2021-22 flu season (figure 4B).

### Changing individual socio-demographic determinants

Changes in responses within demographic groups between both surveys can be investigated by deriving MRP estimates for each socio-demographic group in each year. That is, equation 1 is used to calculate estimates for each relevant demographic cell in each year to obtain estimates. These MRP estimates for each socio-demographic group are presented in figure A.1 and table A.1 for all adults (18+) and figure A.2 and table A.2 for 65-and-overs.

For the entire adult population (18+), every demographic group (with the exception of 65 to 79 year-olds) reported an increase in being offered the flu vaccine (figure A.1 and table A.1). Individuals aged 45 to 54 and 55 to 64 reported very large increases (18.4pp, CrI 15.0 to 20.9 and 36.5pp, CrI 33.5 to 39.3, respectively) due to the expanded seasonal influenza vaccine offer to over 50s in December 2020 [23]. Increases across other socio-demographic groups were broadly uniform at around a 10 percentage point increase (figure A.1 and table A.1). Between the 2020 and 2022 surveys, there were increases in the proportion of respondents reporting that they received a flu vaccine in the previous 12 months among English adults (18+) across all socio-demographic groups with the exception of only a small number of groups (for example 18-24 year-olds and students) whose estimates had a corresponding credible interval that contained zero (figure A.1 and table A.1). While both males and females experienced an increase in influenza vaccine uptake reported in the 2022 survey compared to 2020, females showed a larger increase compared to males (females: 10.4pp, CrI 7.3 to 12.7; males: 7.1pp, CrI 4.0 to 9.7). Regarding changes in willingness to accept a seasonal influenza vaccine among English adults between the 2022 and 2020 surveys, many declines were detected. Asian/Asian British respondents showed a decrease in willingness of 6.8pp, CrI 3.0 to 9.9, with a large fall among Hindu (7.6pp, CrI -14.3, -1.0) and Punjabi-speaking (13.9pp, CrI 1.9 to 25.3) populations sampled (figure A.1 and table A.1). Decreases in willingness to accept a seasonal influenza vaccine in the next 12 months in the 18+ population were also found among both males and females, younger age groups, those with level 1-4 education, as well as respondents who report that they look after the home, work part-time, or reported an ‘other’ employment status. Atheists / agnostics also reported a decrease in willingness between the 2020 and 2022 surveys (figure A.1 and table A.1).

Among the 65-and-over cohort (65+), most socio-demographic groups report a slight decrease in being offered a seasonal influenza vaccine in the previous 12 months between the 2020 and 2022 surveys, but we detect sizeable increases accepting the vaccine if it was offered (figure A.2 and table A.2). White respondents aged 65+ are more likely to report that they have accepted a seasonal influenza vaccine in the last 12 months than Black / Black British respondents in both survey years (White, 2020: 72.8%, CrI 69.8 to 76.0, 2022: 87.0%, CrI 85.1, 88.6; Black/Black British, 2020: 66.9%, CrI 59.2 to 74.8, 2022: 75.1%, Cr: 66.9 to 81.1) as well as a willingness to accept a flu vaccine (White, 2020: 90.9%, CrI 90.0 to 91.8, 2022: 93.0%, CrI 92.2 to 93.9; Black/Black British, 2020: 80.5%, CrI 76.2 to 84.0, 2022: 85.4%, CrI 81.7 to 88.9), despite only small differences in propensity to be offered the vaccine (White, 2020: 89.7%, CrI 88.4 to 91.1, 2022: 87.3%, CrI 86.0 to 88.9; Black/Black British, 2020: 89.0%, CrI 86.3 to 90.9, 2022: 86.0%, CrI 81.4 to 89.5) (figure A.2 and table A.2).

### Predicting regional uptake rates using large-scale surveys and MRP

At the national level, our estimates for receipt of the flu vaccine in the past 12 months agree with uptake rates reported by UK Health Security Agency (UKHSA). Our national-level estimates for the percentage of 65-and-overs who received a seasonal influenza vaccine in the past 12 months given it was offered to them is 72.6% (69.5 to 75.9) in the 2020 survey and 86.6% (84.7 to 88.2) in the 2022 survey (table 2). These figures align well with UKHSA-reported seasonal influenza uptake in this age group of 72.4% and 82.3% in the corresponding 2019-20 and 2021-22 flu seasons [3, 24]. In 2022, 84.4% (82.9 to 85.9) of respondents 65-and-over reported that they would ‘definitely’ take the seasonal influenza vaccine in the next 12 months (interpreted as the 2022-23 winter flu season), while 8.3% (7.6 to 9.0) reported that they were ‘unsure, but leaning towards yes’ (table 2). These values compare to 79.9% reported by UKHSA for the 2022/23 campaign [24]. (We note that to arrive at these estimates for the 65-and-over cohort, estimates were combined across 65 to 79-year-olds and 80 and overs, accounting for respective population sizes.)

A comparison between intent to accept a flu vaccine in 12 months following the 2022 among the 65-and-over cohort and observed uptake for the 2022-23 flu season reveals a strong correlation across the 33 ITL sub-national regions (figure 5). The correlation between observed and predicted values for 65s-and-overs who indicated a positive attitude to taking a flu vaccine in the next 12 months (responding ‘yes, definitely’ or ‘unsure, but leaning towards yes’) is 0.79 (*p <* 0.001) with an average upward bias in predicted values of 13.1pp compared to observed uptake. The correlation between observed and predicted values for those reporting the strongest intent to take a flu vaccine in the next 12 months (that is, responding ‘yes, definitely’ only) is 0.80 (*p <* 0.001) but with a much-reduced upward bias of 4.6pp (figure 5). These observed systematic biases between observed and predicted values may be due to various factors, including an incomplete response rate from GP practices [3], social desirability biases cite, or due to perceived waning importance of the seasonal influenza vaccine as the Covid-19 pandemic subsided.

**Figure 5:**
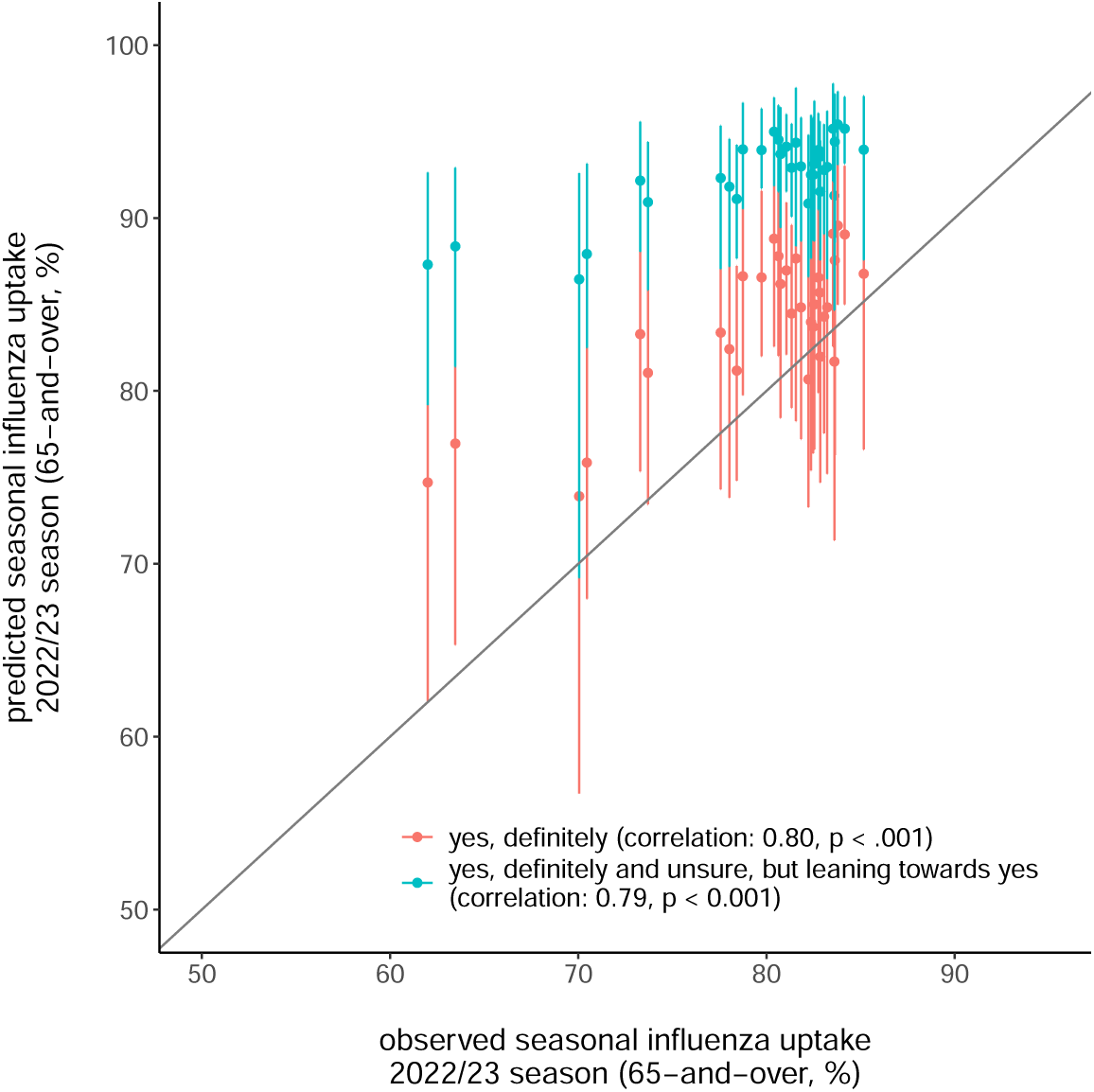
Correlation between MRP estimates of future intent to accept a Covid-19 vaccine in the 2022 survey and observed seasonal influenza uptake in the 2022-23 flu season for all 33 sub-national ITL regions.

## Discussion

The analysis presented in this study sheds light on the evolving landscape of seasonal influenza vaccination behaviours in England amid the Covid-19 pandemic. Our findings highlight ongoing dynamics between public health interventions, socio-demographic factors, and individual attitudes toward vaccination, which we explore in more detail below. Our study also highlights existing socio-demographic gaps in seasonal influenza vaccination trends, as well as revealing a possible early warning signal for confidence in the flu vaccine among Asian and Asian British groups, and notably Punjabi speakers and Hindu respondents.

### Impact of the pandemic and policies

The considerable increase in seasonal influenza vaccine uptake observed between the 2019-20 and 2021-22 flu seasons suggests a positive response to the expanded vaccination efforts in response to the Covid-19 pandemic, including a targeted drive for adults over 50 [23]. The increase in the proportions of those aged 50-and-over both reported being offered a flu vaccine, and accepting the vaccine given that it was offered reflects the successful implementation of an expanded seasonal influenza vaccine [23]. The introduction of novel Covid-19 vaccines and associated communication campaigns likely played a significant role in raising public awareness about the importance of vaccination, potentially contributing to the observed increases. Given the high intent to vaccinate among over 50s in our 2022 survey, the decision to scrap the offer of a seasonal influenza vaccine to over 50s in 2022 [25] may reflect a missed opportunity to secure high adult population vaccination coverage rates and reduce the seasonal burden of influenza in this cohort [26].

The slight decline in vaccine acceptance rates among certain demographic groups, such as individuals aged 18 to 24, as well as among Asian and Asian British individuals, Hindus, and Punjabi speakers, warrants attention. Understanding the factors driving this decrease is crucial for devising interventions to address vaccine hesitancy and ensure equitable vaccine coverage across all groups. Further research is needed to explore the specific barriers and concerns influencing vaccine decision-making in these groups, especially in the context of evolving public health narratives and changing perceptions of vaccination in the wake of the pandemic. Recent evidence in the UK found that Covid-19 vaccine policies themselves may have induced lower vaccine confidence in some groups, though more research is needed to understand how these may translate to non-Covid vaccinations, such as seasonal influenza [10, 27].

### Socio-Demographic disparities

Our analysis reveals persistent socio-demographic disparities in seasonal influenza vaccine uptake, echoing existing literature highlighting inequities in vaccination behaviours across various demographic groups [28, 14]. Notably, individuals from minority ethnic backgrounds, lower education levels, and certain religious affiliations exhibit lower odds of vaccine acceptance, underscoring the need for targeted interventions to address these disparities. Tailored communication strategies and culturally sensitive outreach efforts may be effective in addressing vaccine hesitancy among marginalized communities [29, 30, 31]. Collaborative efforts involving community leaders, healthcare providers, and public health authorities are essential for fostering trust, dispelling misinformation, and promoting vaccine confidence within these populations [29, 30, 32].

### Implications for public health policy

The observed regional variations in vaccine uptake underscore the importance of localised strategies to address specific socio-demographic needs and preferences. Targeted interventions tailored to the socio-economic and cultural context of each region – notably inner London – can enhance vaccine accessibility and uptake, contributing to more effective disease prevention efforts at both national and sub-national levels [33, 34].

The decline in future intent to accept a seasonal influenza vaccine among certain demographic groups highlights the need for sustained efforts to monitor and address evolving attitudes toward vaccination. Continued surveillance of vaccine sentiment and behaviour is essential for informing evidence-based policy decisions and adapting public health interventions to effectively address emerging challenges [35]. Our study has shown that future rates of uptake can be reliably predicted from a combination of large-scale data collection re-weighted against individual census records. Such a prediction and monitoring tool could be used to identify emerging confidence issues, ensuring rapid interventions sensitive to local community needs.

### Limitations and future directions

While this study provides insights into the factors influencing seasonal influenza vaccine uptake in England, several limitations should be acknowledged. The reliance on survey data introduces the potential for response bias and self-reporting errors, which may influence the accuracy of vaccine uptake estimates. Additionally, the cross-sectional nature of the study precludes causal inference and temporal analysis of vaccine trends over longer periods.

In conclusion, addressing socio-demographic disparities and promoting vaccine acceptance remain critical priorities for public health efforts to enhance seasonal influenza vaccination coverage in England. By employing multifaceted approaches that integrate community engagement, targeted communication, and evidence-based policy interventions, policymakers and healthcare stakeholders can strive toward achieving equitable access to vaccination and safeguarding population health in the post-pandemic era.

## Data Availability

Study data is available upon request from the corresponding author.

## Appendix A. Appendix

**Figure A.1:**
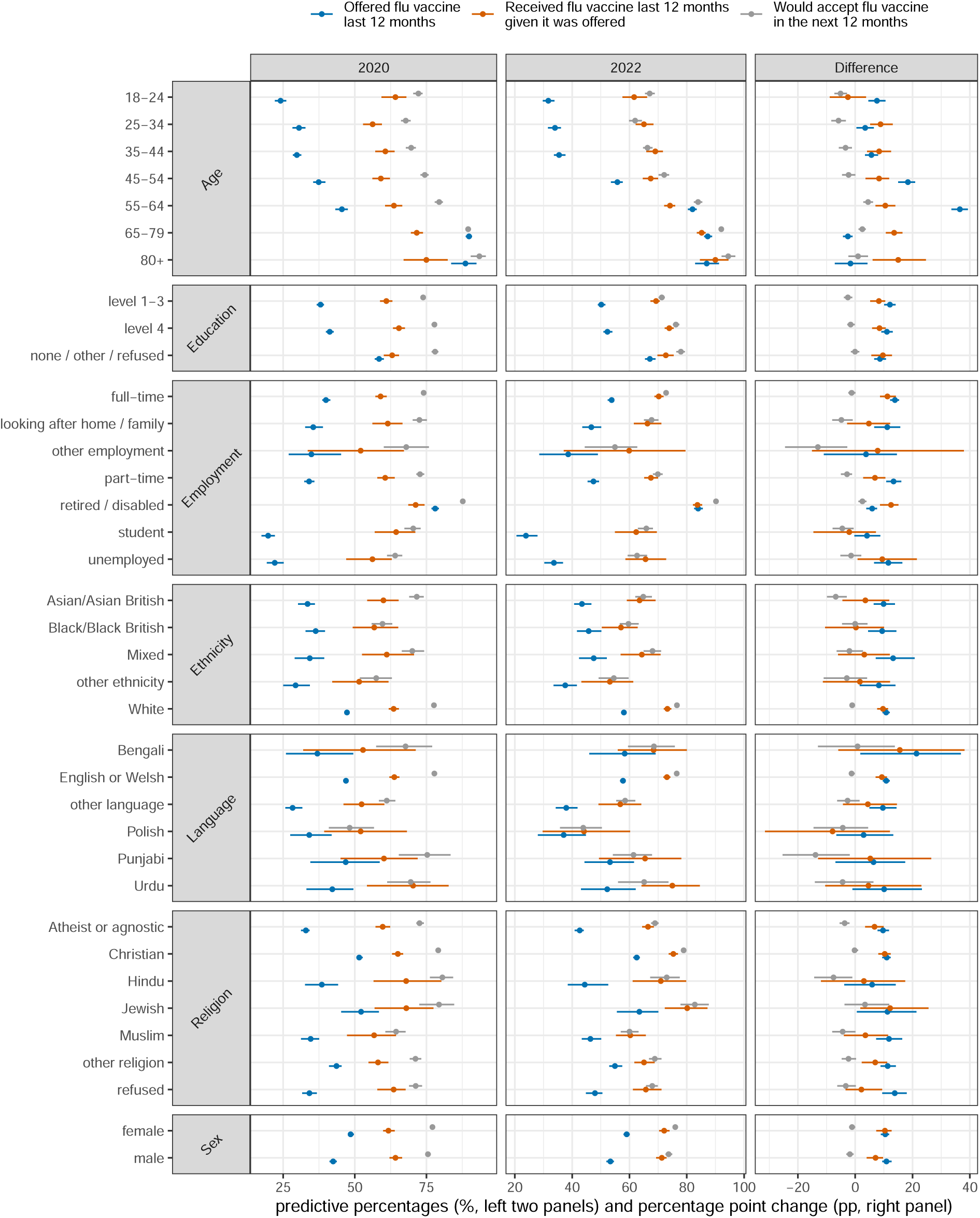
MRP estimates to each response variable for each socio-demographic group for the adult population (18+) MRP estimates for each question and socio-demographic group in 2020 and 2022 (left and centre panels) and the percentage point change in responses to each question between 2020 and 2022 (right panel), where a positive percentage point change signifies an increase between 2020 and 2022.

**Figure A.2:**
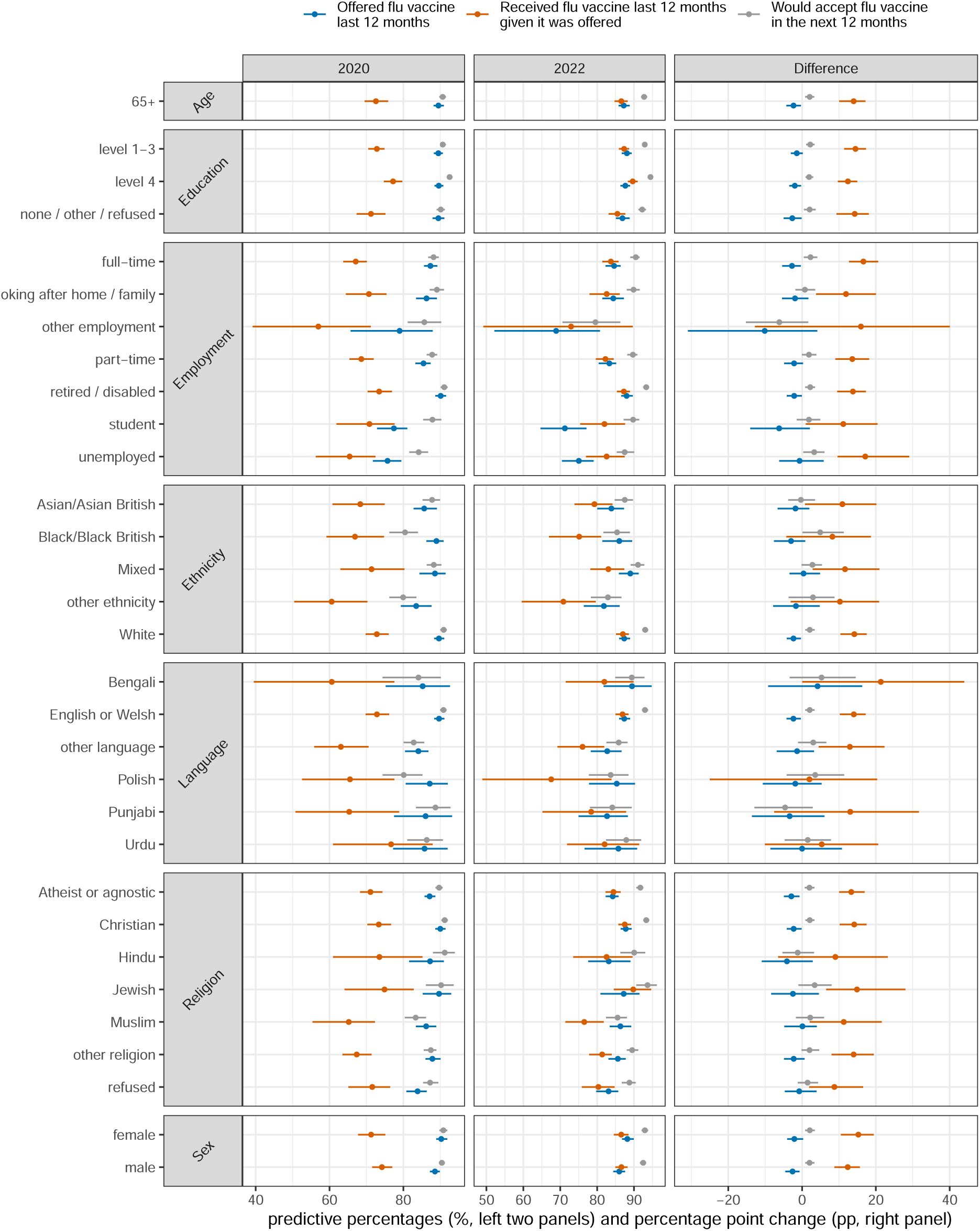
MRP estimates to each response variable for each socio-demographic group for the 65-and- over population (65+) MRP estimates for each question and socio-demographic group in 2020 and 2022 (left and centre panels) and the percentage point change in responses to each question between 2020 and 2022 (right panel), where a positive percentage point change signifies an increase between 2020 and 2022.

**Table A.1:**
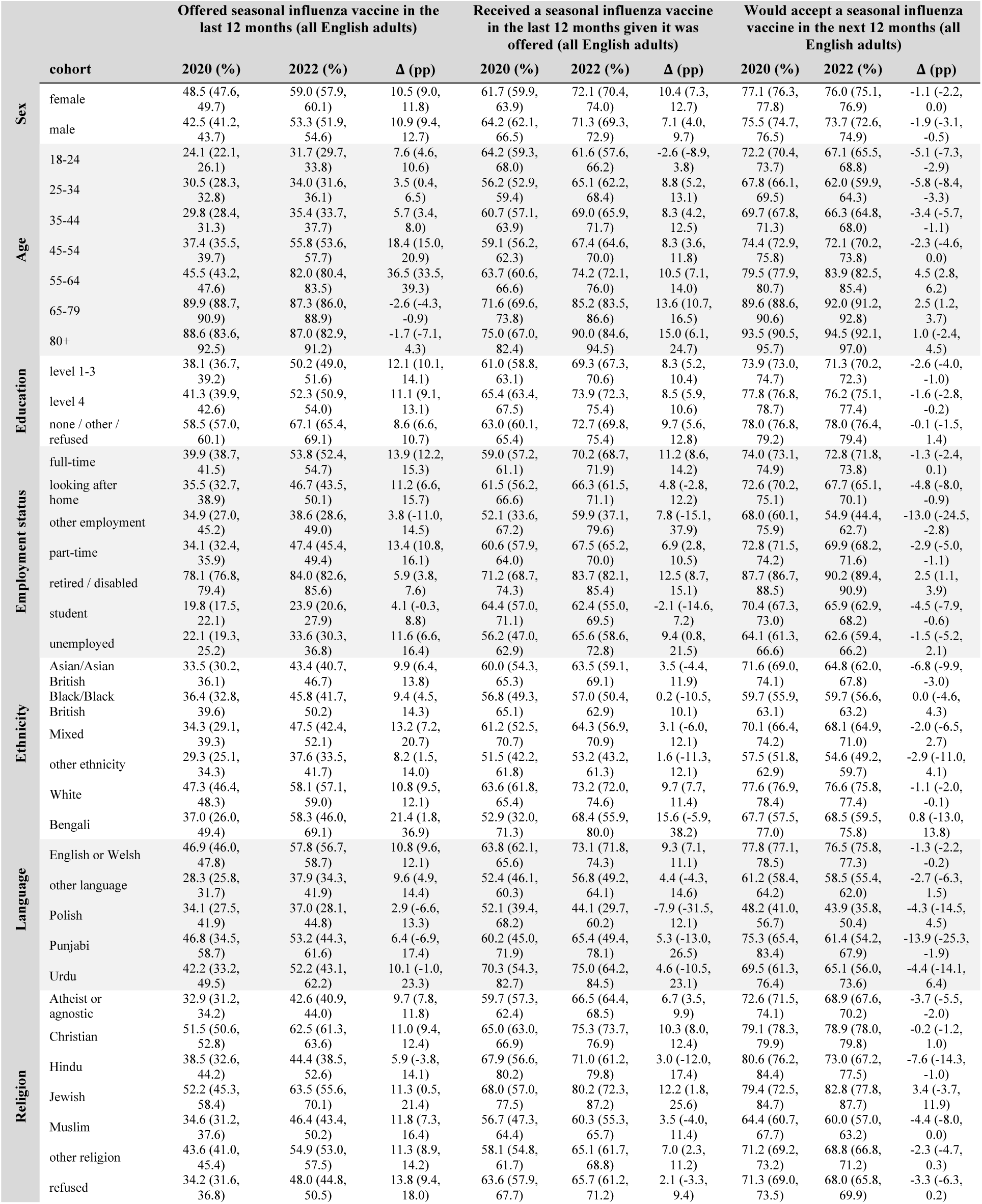
MRP estimates to each response variable for each socio-demographic group for the adult population (18+) in tabular form. MRP estimates for each question and socio-demographic group in 2020 and 2022 and the percentage point change in responses to each question between 2020 and 2022, where a positive percentage point change signifies an increase between 2020 and 2022. The raw data in this table is plotted in figure A.1.

**Table A.2:**
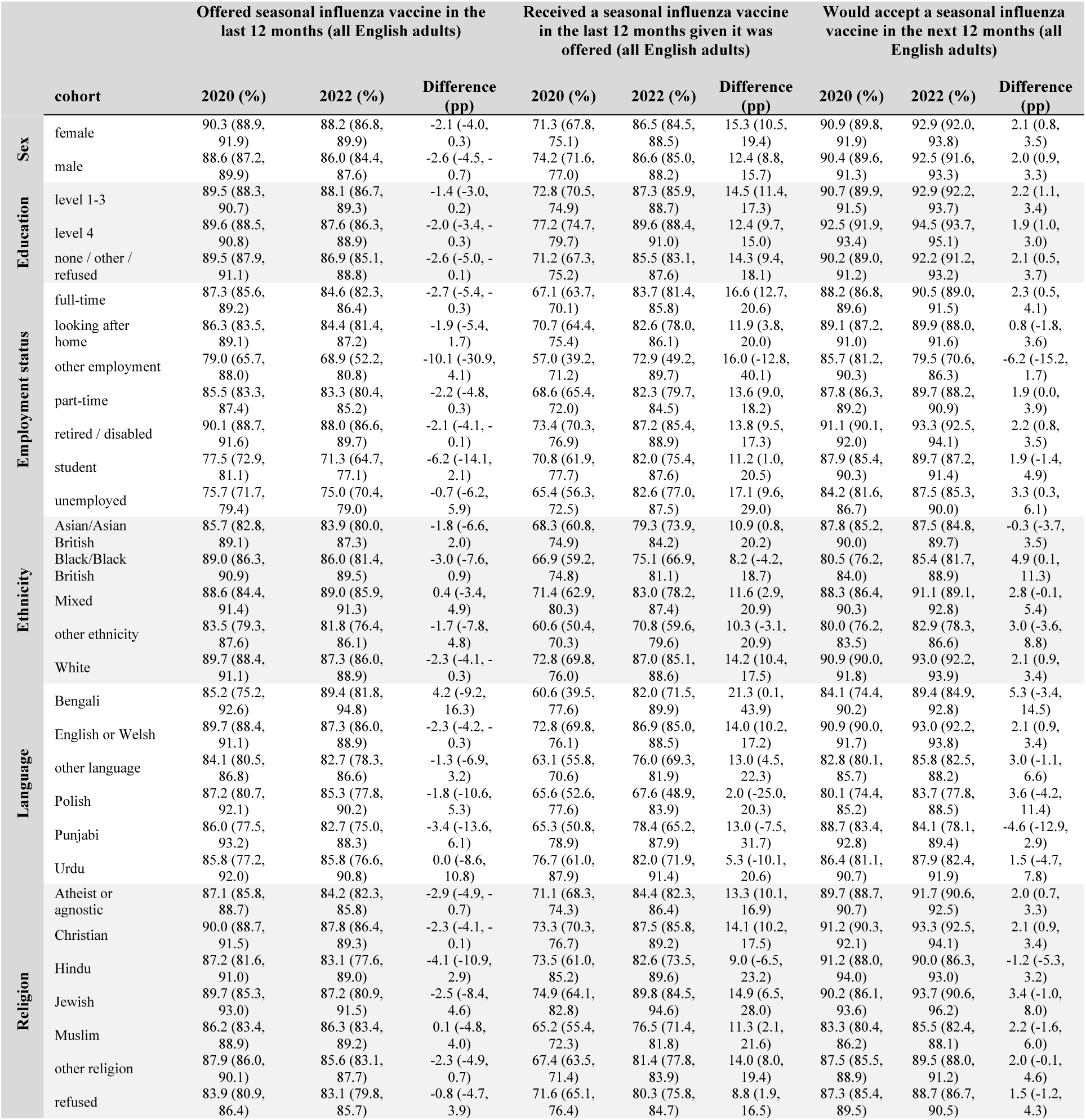
MRP estimates to each response variable for each socio-demographic group for the 65-and- over population (65+) in tabular form. MRP estimates for each question and socio-demographic group in 2020 and 2022 and the percentage point change in responses to each question between 2020 and 2022, where a positive percentage point change signifies an increase between 2020 and 2022. The raw data in this table is plotted in figure A.2

